# Reactogenicity and immunogenicity against MPXV of the intradermal administration of Modified Vaccinia Ankara compared to the standard subcutaneous route

**DOI:** 10.1101/2024.09.17.24313609

**Authors:** Valentina Mazzotta, Pierluca Piselli, Alessandro Cozzi Lepri, Giulia Matusali, Eleonora Cimini, Rozenn Esvan, Francesca Colavita, Roberta Gagliardini, Stefania Notari, Alessandra Oliva, Silvia Meschi, Rita Casetti, Giulia Micheli, Licia Bordi, Alessandro Giacinta, Germana Grassi, Saba Gebremeskel Tekle, Claudia Cimaglia, Jessica Paulicelli, Alessandro Caioli, Paola Gallì, Giulia Del Duca, Miriam Lichtner, Loredana Sarmati, Enrica Tamburrini, Claudio Mastroianni, Alessandra Latini, Paolo Faccendini, Carla Fontana, Emauele Nicastri, Andrea Siddu, Alessandra Barca, Francesco Vaia, Enrico Girardi, Fabrizio Maggi, Andrea Antinori, MPOX Vaccine Lazio Study Group

## Abstract

The recent resurgence of Mpox in central Africa has been declared again a Public Health Emergency of International Concern (PHEIC) requiring coordinated international responses. Vaccination is a priority to expand protection and enhance control strategies, but the vaccine’s need exceeds the currently available doses. Intradermal administration of one-fifth of the standard Modified-Vaccinia-Ankara (MVA-BN) dose was temporarily authorized during the 2022 PHEIC. Studies conducted before 2022 provided evidence about the humoral response against the Vaccinia virus (VACV) after vaccination but not against the Mpox virus (MPXV). Moreover, no data are available on the T-cell response elicited by MVA-BN administered subcutaneously or intradermally. Here, we compare the two vaccine administration routes according to reactogenicity and immunogenicity based on data from 943 vaccine recipients during the 2022 vaccination campaign in Rome, Italy. We found that the intradermal route elicited slightly higher titers of MPXV-specific IgG and nAbs than the subcutaneous one. At the same time, no differences in cellular response were detected. MVA-BN was globally well tolerated despite higher reactogenicity for the intradermal than the subcutaneous route, especially for the reactions at the local injection site. The intradermal dose-sparing strategy was proven safe and immunogenic and would make vaccination available to more people.

## Introduction

As of May 2022, an unexpected mpox epidemic caused by the clade IIb spread in 122 countries with a global case count higher than 99.000^1^. The rapid increase in confirmed cases led to the subsequent declaration of mpox as a public health emergency of international concern (PHEIC) by the World Health Organization (WHO)^2^. Therefore, the WHO recommended vaccination for high-risk people and other mitigation strategies to control the epidemic^3^.

Licensed vaccines against mpox consist of two doses of the third-generation replication-deficient modified vaccinia Ankara produced by Bavarian Nordic (MVA-BN)^4,5^. The standard route of administration of MVA-BN is subcutaneous (SC), with two doses delivered at least 28 days apart. However, a randomized clinical trial (RCT) compared the safety and immunogenicity of the standard SC formulation (dose and route) of MVA-BN with the intradermal (ID) administration of one-fifth of the standard SC dose^6^. Although the proportion of local reactions was significantly higher after ID inoculation than SC, the ID route was considered non-inferior to the SC route regarding neutralization against vaccinia virus, and it was purposed as a valid alternative in the event of an emergency requiring more doses available^7^.

In the summer of 2022, it became necessary to rapidly vaccinate as many high-risk people as possible to contain the epidemic ^8^. Therefore, due to the limited availability of MVA-BN worldwide, the European Medicine Agency and Food and Drugs Administration authorized using the intradermal route of administration with a reduced dose to extend vaccination ^9,10^.

Data from large-scale administration of MVA-BN published by Bavarian Nordic confirmed the overall safety of the vaccine with an increased frequency of syncopal events after ID administration^11^. Recently, Frey et al. reported data from a phase-2 open-label trial comparing two 2-dose ID regimens of MVA-BN (one-tenth and one-fifth) with the standard dose SC regimen. MVA-BN administrated ID at fractional doses was safe, and the vaccination with one-fifth (but not one-tenth) dose demonstrated non-inferior immunogenicity by vaccinia virus (Western Reserve strain) plaque reduction neutralization test (PRNT) after 43 days^12^. However, even though similar clinical effectiveness of the two different vaccination schedules against mpox was reported from 2022 real-world data^13^, comparative data on the immunogenicity of ID route compared to SC are lacking and, those available are not informative on the humoral response specifically against mpox, nor on the T-cell response.

Although the massive vaccination campaigns during the MPXV Clade IIb outbreak must have helped to control the epidemic in high-income countries^14^, many outbreaks were constantly observed in endemic areas during the last year, especially in South Africa, where Clade IIb of mpox virus (MPXV) was still detected^15^ and in the Democratic Republic of Congo, where the Clade I had acquired the capability of human-to-human transmission through the sexual route^16^, and it has spread in some neighboring countries where mpox cases have never been reported before (e.g Burundi, Rwanda, and Uganda)^17^. Furthermore, this “new sexually transmitted clade”, named Clade Ib, is particularly worrisome because of its lethality (nearly 3%)^18^, leading WHO to renew the declaration of mpox as a PHEIC on the 14^th^ of August 2024^17^.

Vaccination is recommended as one of the main tools to control this new increase in mpox cases. WHO and the African Centers for Disease Control and Prevention (CDC) are producing efforts to step up the vaccination campaign in low-income countries with limited access. In front of the 10 million doses African CDC estimates are needed to contain the current outbreak, the negotiations with Bavarian Nordic aim to obtain around 200.000 doses of MVA-BN^19^. In this setting, enhancing knowledge about the reactogenicity and immunogenicity against MPXV of the intradermal dose-sparing strategy compared to the standard subcutaneous one in mpox high-risk people in the real-world setting is crucial to address an efficient and cost-effective implementation of the arising mpox vaccination campaign.

Here, we report observational data from a data set on the non-randomized comparison on safety and reactogenicity, and immunogenicity (neutralization and T-cell response) specifically directed against the MPXV target between the two vaccination schedules of MVA-BN (Jynneos) delivered in the vaccination campaign during the 2022-2023 mpox outbreak in Italy.

## Methods

### Patients’ enrolment

In the Lazio Region of Italy mpox vaccination campaign started on August 8th, 2022, and took place in a hospital setting at the Lazzaro Spallanzani National Institute for Infectious Diseases in Rome, which was identified as the only vaccination center in the entire region.

According to the recommendations of the Ministry of Health^20^, MVA-BN was administered as pre-exposure prophylaxis to a target population, including laboratory personnel with possible direct exposure to orthopoxviruses (OPXV) and high-risk gay-bisexual-men who have sex with men (GBMSM), defined as individuals reporting a recent history of sexually transmitted infections (STIs), multiple sexual partners or participation in group sex events, sexual encounters in clubs/cruising/saunas or sexual acts associated with the use of Chemical drugs (Chemsex). For individuals who had never received the smallpox vaccine (vaccine-naïve or non-primed), the vaccination schedule consisted of a two-dose cycle with a 28-day interval between each dose, while individuals who had received the smallpox vaccine in the past (vaccine-experienced or primed) were administered a single-dose cycle. Of note, in Italy, the smallpox vaccination campaign was stopped in 1977 and officially abrogated in 1981 ^21^.

The MVA-BN first dose was administered subcutaneously during the first two weeks of the vaccination campaign, after which the intradermal route was adopted, following the ministerial indications^22^.

For the same reason, the vaccine was administered exclusively intradermically in those receiving a second dose. A prospective observational cohort was integrated into the framework of this vaccination campaign.

### Study protocol

The protocol for the study named Mpox-Vac (“Studio prospettico osservazionale per monitorare aspetti relativi alla sicurezza, all’efficacia, all’immunogenicità e all’accettabilità della vaccinazione anti Monkeypox con vaccino MVA-BN (JYNNEOS) in persone ad alto rischio”) was approved by the INMI Lazzaro Spallanzani Ethical Committee (approval number 41z, Register of Non-Covid Trials 2022). The study protocol was previously described in detail ^23^.

Briefly, all subjects eligible for mpox vaccination according to the ministerial guidelines and who signed a written informed consent were enrolled in the study. Laboratory personnel were excluded from the analysis. At baseline (when receiving the first MVA-BN dose, T1), subjects were evaluated for demographic and behavioural characteristics linked to mpox exposure. Information regarding HIV status, CD4 count, HIV pre-exposure prophylaxis (PrEP) uptake and any history of previous STIs was collected. Other time points were scheduled at the administration of the second dose (T2) and one month after the completion of the cycle (T3). For the vaccine-experienced individuals (primed) who received a single-dose schedule as a complete vaccination cycle, T2 was the time point corresponding to one month after vaccination completion.

### Assessment of adverse reactions

As part of the protocol, participants were delivered a paper symptoms diary after each vaccine dose (T1 and T2) to collect self-reported adverse effects following immunization (AEFIs) for 28 consecutive days. Participants returned the completed diaries at the next time point.

Participants were able to report the presence of systemic symptoms (S-AEFIs) classified as fatigue, muscle pain, headache, gastrointestinal effects, and chills, and local injection site symptoms (LIS-AEFIs), such as redness, induration, and pain. AEFIs were graded by the vaccinees as absent (grade 0), mild (1), moderate (2), or severe (3). Here, we report the results regarding erythema and induration as recalculated with the use of the current FDA Toxicity Grading Scale for Healthy Adult and Adolescent Volunteers Enrolled in Preventive Vaccine Clinical Trials. According to that scale, a diameter of 25 to 50 mm indicates a mild reaction, 51 to 100 mm is a moderate reaction, and more than 100 mm is a severe reaction^24^.

### Assessment of immunogenicity

In a specific subgroup of participants for whom blood samples collected at each time point were available, the assessment of the early humoral and cellular immune response was performed.

### MPXV-specific IgG and neutralization assays

Specific anti-MPXV immune response was evaluated by measuring MPXV-specific IgGs and neutralizing antibodies in the serum as previously described^25^. The presence of anti-MPXV IgGs was assessed on immunofluorescence slides in-house prepared with Vero E6 cells (ATCC) infected with an MPXV isolated from the skin lesion of a patient infected with MPXV during the 2022 outbreak (GenBank: ON745215.1, referred to the clinical sample). Serum samples were tested with a starting dilution of 1:20, and serial two-fold dilutions were performed to determine anti-MPXV IgG titer. MPXV-specific neutralizing antibodies (nAbs) were measured by 50% plaque-reduction-neutralization test (PRNT_50_) with a starting dilution of 1:10. Specifically, serum samples were heat-inactivated at 56 °C for 30 min and titrated in duplicate in 4 four-fold serial dilutions. Each serum dilution was added to the same volume (1:1) of a solution containing 100 TCID50 MPXV isolate (GenBank: ON745215.1, referred to the clinical sample) and incubated at 37 °C for 2 h. Subconfluent Vero E6 cells were infected with virus/serum mixtures and incubated at 37 °C and 5% CO_2_. After 5 days, the supernatant was carefully discarded, and a crystal violet solution (Diapath S.P.A.) containing 10% formaldehyde (Sigma-Aldrich) was added for 30 min, then cells were washed with phosphate-buffered saline (PBS, 1X; Sigma-Aldrich). Using the Cytation 5 reader (Biotek), the number of plaques was counted. The neutralizing titers were estimated by measuring the plaques number reduction as compared to the control virus wells. The highest serum dilution showing at least 50% of the plaque number reduction was indicated as the 50% neutralization titer (PRNT_50_). Each test included serum control (1:10 dilution of each sample tested without virus), cell control (Vero E6 cells alone), and virus control (100 TCID_50_ MPXV in octuplicate).

### PBMC isolation

Using Ficoll density gradient centrifugation (Pancoll human, PAN Biotech) methodperipheral blood mononuclear cells (PBMC) were isolated, frozen in FBS (Fetal Bovine Serum, Gibco, USA) added of 10% of DMSO (Merck Life sciences, Milan, Italy) at vapors of liquid nitrogen for further experiments.

### Elispot assay

The frequency of T-cell-specific responses to the MVA-BN vaccine was assessed by Interferon-γ ELISpot assay. Briefly, PBMC were thawed and suspended in a complete medium [RPMI-1640 added of 10% FBS, 1% L-glutamine, and 1% penicillin/streptomycin (Euroclone S.p.A, Italy)]. Live PBMC were counted by Trypan blue exclusion, plated at 3×10^5^ cells per well in ELISpot plates (Human IFN-γ ELISpot plus kit; Mabtech, Nacka Strand, Sweden), and stimulated for 20 h with MOI 1 of the MVA-BN vaccine suspension [JYNNEOS (Smallpox and Monkeypox Vaccine, Live, non-replicating)] and αCD28/αCD49d (1 μg/ml, BD Biosciences) at 37 °C (5% CO_2_), using T cell superantigen (SEB 200 nM, Sigma) as positive control. At the end of the of incubation, the ELISpot assay was developed following manufacturer’s instructions. Results are expressed as spot-forming cells per 10^6^ PBMCs (SFC/10^6^ PBMCs) in stimulating cultures after subtracting the background (unstimulated culture).

### Statistical analysis

Demographic and epidemiological characteristics of the patients are presented both for the overall sample population and for each route of administration. Continuous variables were described using median and Interquartile Range (IQR), while categorical variables were summarized using absolute and relative (percentage) frequencies.

The main characteristics of participants according to the route of administration were compared using the Chi-square test or Fisher exact test when appropriate for qualitative variables and the Mann-Whitney test for numerical variables. We then used collected diaries to calculate the prevalence of reported AEFIs according to the route of administration, as well as the duration and maximum level of severity ever experienced over the 28 days following the first dose of vaccination for each S-AEFI and LIS-AEFI, according to the above-mentioned grading (none to severe).

The raw proportions of participants whose maximum reported AEFIs was none, mild, moderate or severe by route of administration groups are also shown. We also computed the Odds ratio (OR) of the maximum severity level of AEFIs experienced by the administration route using multinomial logistic regression models, both univariable and after adjusting for age and HIV status. In these models, the never-reported AEFIs category was chosen as the reference group, and the estimated ORs show the risk of reporting mild, moderate or severe AEFIs as the maximum level ever experienced according to the administration route. For a proportion of participants, the diary data were censored at day six, so we have also performed a sensitivity analysis restricting to only the level of AEFI severity ever experienced over the six days following the first vaccination dose for everybody.

As a second continuous outcome, we calculated the average number of days in which participants experienced each of the 4 levels of symptoms over the 28 days past the first dose of the MVA-BN vaccine. We then compared the average duration in days of any systemic or local reactions by route of administration by means of an unpaired t-test. Also, in this analysis, the comparison was performed by analyzing the reporting of any grade of reaction (from mild to severe, grade 1 to 3) or moderate to severe (2 or 3).

Similarly to what was done for the categorical outcome, we controlled for the potential confounding effect of age and HIV status using a counterfactual linear regression model and provided an estimate of the average treatment effect (ATE) associated with the administration route. Of note, the aim of this analysis was to evaluate what would have been the duration of any adverse response to the MVA-BN vaccine had everybody in the sample received the ID vs. had everybody received the SC route of administration instead. We used a doubly robust method (using augmented inverse probability weighting-AIPW) to obtain estimates which are robust against misspecification of either the propensity model or the outcome model. Both the propensity and outcome models included HIV and age as confounding variables. For the outcome model, we also used the saturated model, including the interaction parameter between exposure and HlV status (results were similar, not shown).

Finally, the same approach to analysis (a marginal model and the calculation of the ATE after controlling for HIV and age) was conducted in a subset of 171 (18%) vaccinated subjects for whom stored samples were available and analyzed to compare the overall increase in the average levels (on a log_2_ scale) of IgG and nAb as well as ELISPOT response 1 month after vaccination according to route of administration.

## Results

### Characteristics of the study population

Between August 8th and December 31st, 2022, 3,296 individuals received at least one dose of MVA-BN. Of those, 943 (28.6%) agreed to be enrolled in the study and completed the symptom-reporting daily diary covering their experience within 28 days from the first dose administration. Among the 943 individuals included in the analysis, 225 (23.9%) received the first dose via the SC route, of whom 50 as a single dose because they were vaccine-experienced (22.2% of this group), and 718 (76.1%) via the ID route; of this latter group 523 (73%) received only the first of the two doses scheduled for the previously unvaccinated.

All were male, and the majority (90.9%) self-identified as MSM. Overall, the median age was 44 years (IQR 36-51), 43 (36-48) years in the SC group, and 45 (38-52) years in the ID, respectively (p=0.15). Regarding other characteristics, 167 participants (17.7%) were on PrEP, 227 (24.1%) reported at least one STI diagnosed within the previous year, and 261 subjects (27.7%) were people living with HIV (PLWH), all on highly active antiretroviral therapy. HIV infection was more prevalent in the SC (35%) vs. the ID group (25%, p=0.0004). In those with HIV, CD4-cell count was lower than 200 cells/μL only in 10 participants (3.8%), while it was higher than 500 cells/μL in 211 (80.8%), with no evidence for a difference between the two groups (p=0.232). The two groups differed significantly also according to the use of PrEP (25.8% vs. 15.2%, for SC and ID groups respectively, p<0.001) and in the proportion of participants who reported one or more comorbidities (0% vs. 8.2%, for SC and ID group, respectively, p<0.001), while there was no evidence for a difference in other examined factors (i.e. sexual orientation, history of STI other than HIV, CD4 counts for those with HIV, and history of smallpox vaccination. The main characteristics of the study population according to the administration route are reported in more detail in Table 1. The crude proportions of participants reporting various grades of the evaluated adverse effect are reported in Figure 1 and Table 2. The grade and duration of S-AEFIs and LIS-AEFIs within 28 days from vaccination according to the route of vaccination are summarized in Supplementary Figure 1.

**Figure 1.**
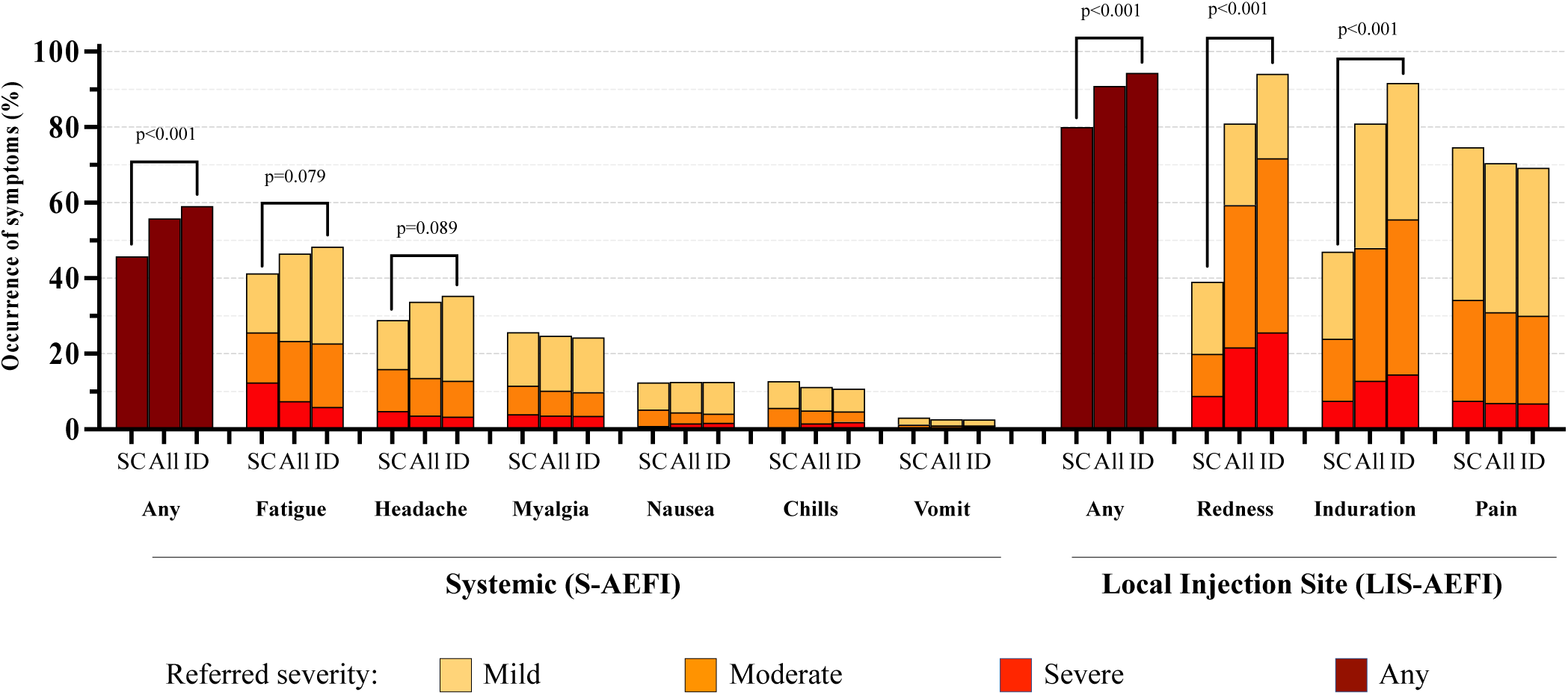
Distribution of reactivity and referred severity of Systemic and Local Injection Site Adverse Effect Following Immunisation (S-AEFIs and LIS-AEFIs) with the first dose of MVA-BN Vaccine within 28-days from vaccination according to route of administration [All: N=943; Sub-cutaneous (SC): N=225; Intra-dermal (ID): N=718]. p<0.10 are shown; p values refer to the difference in the proportion of any grade symptom between the SC and ID groups.

**Table 1.**
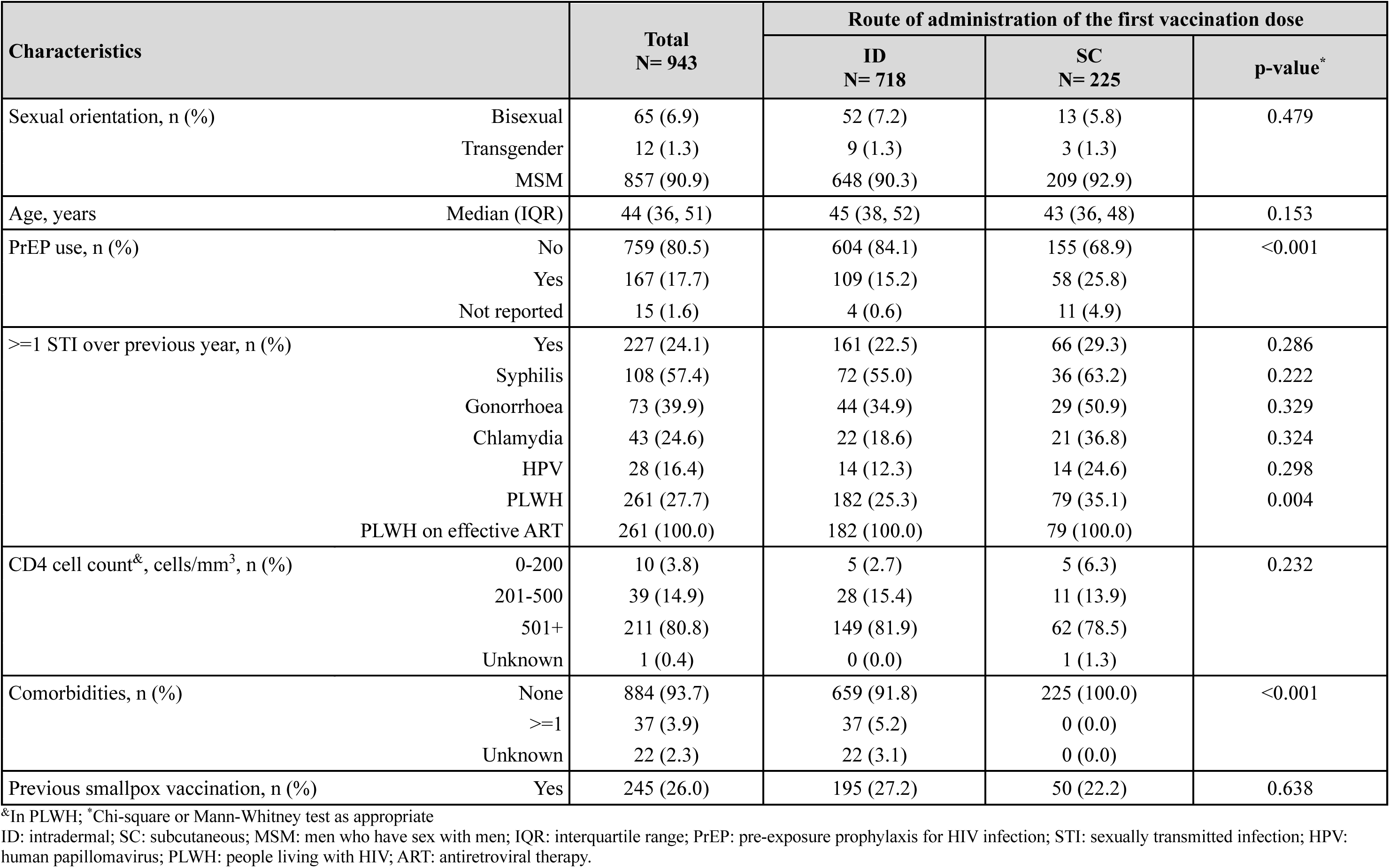
Main characteristics by route of administration of the first MVA-BN dose.

**Table 2.**
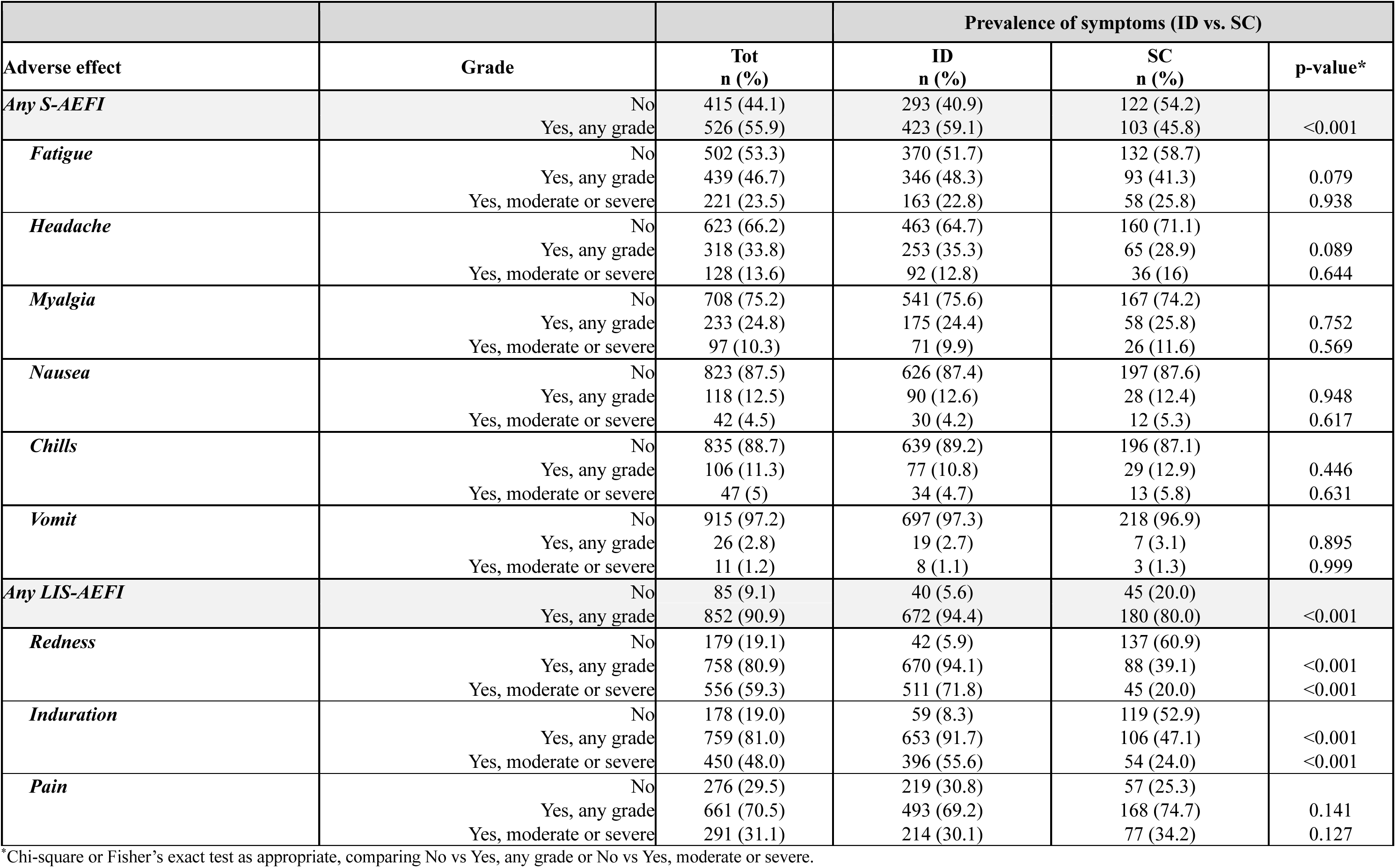
Table of prevalence of Systemic and Local Adverse Effects Following Immunisation (S-AEFIs and L-AEFIs) with MVA-BN Vaccine.

### Systemic reactions

No serious adverse events were observed over the 28-day follow-up. Overall systemic reactions occurred in 526 (55.9%) participants, with a higher proportion observed in the ID group compared to the SC group (59.1% vs 45.8%, p<0.001). The most common S-AEFIs were fatigue and headache, occurring in 46.7% and 33.8% of participants, which tended to be slightly higher in the ID group (p=0.08 and p=0.09, respectively); however, when considering only moderate or severe grades, there was no evidence for a difference between the two groups.

After adjusting for age and HIV status in a multinomial regression model, we found evidence that participants in the ID group had an increased risk of developing mild-grade headaches (2.91; 95% Confidence Interval, 95% CI: 1.23,6.89; p=0.045) compared to the SC group (Table 3). Results were similar when we restricted the analysis only to the first 6 days of the diary (OR=2.66, 95% CI:1.13-6.27, p=0.07), suggesting that most of the difference is likely to occur early after vaccination (Supplementary Table 1).

**Table 3.** Prevalence and risk of developing different grades of Systemic and Local Adverse Effects Following Immunisation (S-AEFIs and L-AEFIs) with MVA-BN Vaccine from fitting a multinomial logistic regression according to administration route - intradermal (ID) vs subcutaneous (SC).

|  |  | Prevalence of S-AEFIs and OR (ID vs. SC) |  |  |  |  |  |
| --- | --- | --- | --- | --- | --- | --- | --- |
| Systemic symptoms | max severity | ID<br>n (%) | SC<br>n (%) | Unadjusted OR (95% CI)<br>ID vs. SC | p-value | Adjusted* OR (95% CI)<br>ID vs. SC | p-value |
| Fatigue | None | 370 (51.7) | 132 (58.7) | 1 | <0.001 | 1 | 0.673 |
|  | Mild | 183 (25.6) | 35 (15.6) | 1.87 (1.23, 2.82) |  | 0.95 (0.40, 2.24) |  |
|  | Moderate | 120 (16.8) | 30 (13.3) | 1.43 (0.91, 2.23) |  | 1.67 (0.66, 4.26) |  |
|  | Severe | 43 (6.0) | 28 (12.4) | 0.55 (0.33, 0.92) |  | 1.54 (0.49, 4.81) |  |
| Headache | None | 463 (64.7) | 160 (71.1) | 1 | 0.011 | 1 | 0.045 |
|  | Mild | 161 (22.5) | 29 (12.9) | 1.92 (1.24, 2.96) |  | 2.91 (1.23, 6.89) |  |
|  | Moderate | 68 (9.5) | 25 (11.1) | 0.94 (0.57, 1.54) |  | 1.37 (0.44, 4.27) |  |
|  | Severe | 24 (3.4) | 11 (4.9) | 0.75 (0.36, 1.57) |  | 7.17 (0.74, 69.81) |  |
| Myalgia | None | 541 (75.6) | 167 (74.2) | 1 | 0.915 | 1 | 0.675 |
|  | Mild | 104 (14.5) | 32 (14.2) | 1.00 (0.65, 1.55) |  | 1.04 (0.39, 2.78) |  |
|  | Moderate | 45 (6.3) | 17 (7.6) | 0.82 (0.46, 1.47) |  | 1.27 (0.37, 4.32) |  |
|  | Severe | 26 (3.6) | 9 (4.0) | 0.89 (0.41, 1.94) |  | 2.47 (0.57, 10.68) |  |
| Nausea | None | 626 (87.4) | 197 (87.6) | 1 | 0.294 | 1 | 0.723 |
|  | Mild | 60 (8.4) | 16 (7.1) | 1.18 (0.66, 2.10) |  | 1.79 (0.50, 6.46) |  |
|  | Moderate | 17 (2.4) | 10 (4.4) | 0.53 (0.24, 1.19) |  | 0.55 (0.10, 3.13) |  |
|  | Severe | 13 (1.8) | 2 (0.9) | Nd |  | Nd |  |
| Chills | None | 639 (89.2) | 196 (87.1) | 1 | 0.089 | 1 | 0.698 |
|  | Mild | 43 (6.0) | 16 (7.1) | 0.82 (0.45, 1.50) |  | 1.13 (0.37, 3.50) |  |
|  | Moderate | 20 (2.8) | 12 (5.3) | 0.51 (0.25, 1.06) |  | 0.27 (0.03, 2.48) |  |
|  | Severe | 14 (2.0) | 1 (0.4) | Nd |  | Nd |  |
| Vomit | None | 697 (97.3) | 218 (96.9) | 1 | 0.195 | 1 | 0.748 |
|  | Mild | 11 (1.5) | 4 (1.8) | 0.86 (0.27, 2.73) |  | 2.68 (0.21, 33.94) |  |
|  | Moderate | 3 (0.4) | 3 (1.3) | Nd |  | Nd |  |
|  | Severe | 5 (0.7) | 0 (0.0) | Nd |  | Nd |  |
|  |  | Prevalence of L-AEFIs and OR (ID vs. SC) |  |  |  |  |  |
| Local symptoms | max severity | ID<br>n (%) | SC<br>n (%) | Unadjusted OR (95% CI)<br>ID vs. SC | p-value | Adjusted* OR (95% CI)<br>ID vs. SC | p-value |
| Redness | None | 42 (5.9) | 137 (60.9) | 1 | 0.089 | 1 | <0.001 |
|  | Mild | 159 (22.3) | 43 (19.1) | 12.06 (7.44, 19.54) |  | 26.87 (6.87, 105.1) |  |
|  | Moderate | 328 (46.1) | 25 (11.1) | 42.80 (25.10, 72.98) |  | 65.91 (16.39, 265.1) |  |
|  | Severe | 183 (25.7) | 20 (8.9) | 29.85 (16.77, 53.13) |  | 48.44 (11.71, 200.4) |  |
| **Induration** | None | 59 (8.3) | 119 (52.9) | 1 | <0.001 | 1 | <0.001 |
|  | Mild | 257 (36.1) | 52 (23.1) | 9.97 (6.47, 15.35) |  | 12.08 (4.44, 32.89) |  |
|  | Moderate | 292 (41.0) | 37 (16.4) | 15.92 (10.02, 25.29) |  | 14.92 (5.03, 44.22) |  |
|  | Severe | 104 (14.6) | 17 (7.6) | 12.34 (6.77, 22.49) |  | 16.28 (4.73, 55.98) |  |
| **Pain** | None | 219 (30.8) | 57 (25.3) | 1 | 0.425 | 1 | 0.002 |
|  | Mild | 279 (39.2) | 91 (40.4) | 0.80 (0.55, 1.16) |  | 0.36 (0.16, 0.81) |  |
|  | Moderate | 165 (23.2) | 60 (26.7) | 0.72 (0.47, 1.08) |  | 0.14 (0.05, 0.39) |  |
|  | Severe | 49 (6.9) | 17 (7.6) | 0.75 (0.40, 1.40) |  | 0.57 (0.15, 2.18) |  |

\* Adjusted for age and HIV status.
95% CI: 95% Confidence Interval
Nd: not determined

Systemic reactions were of short duration, 3.7 days on average for any grade and any type of S-AEFI, with no evidence for a difference in symptom duration between the ID and SC groups in the unadjusted analysis and after controlling for HIV and age (Table 4).

**Table 4.** Contrasts of the duration of Systemic and Local Adverse Effects Following Immunisation (S-AEFIs and L-AEFIs) with MVA-BN Vaccine (days 1-28 after 1st dose) and ATE^&^ from fitting a linear regression model.

|  | Potential average duration of systemic symptoms (days) over 1-28 days after the 1<sup>st</sup> vaccine dose |  |  |  |
| --- | --- | --- | --- | --- |
|  | Mean in ID<br>(95% CI) | Mean in SC<br>(95% CI) | ATE<sup>*</sup> (95% CI)<br>ID vs. SC | p-value |
| ***S-AEFIs*** |  |  |  |  |
| *Fatigue - any* | 3.70 (2.63, 4.77) | 3.46 (2.21, 4.72) | 0.24 (-1.36, 1.84) | 0.7720 |
| *Fatigue - moderate or severe* | 2.95 (1.26, 4.63) | 3.03 (1.07, 4.98) | -0.08 (-2.52, 2.36) | 0.9488 |
| *Headache - any* | 2.20 (1.53, 2.87) | 2.51 (1.81, 3.20) | -0.30 (-1.24, 0.63) | 0.5269 |
| *Headache - moderate or severe* | 2.53 (0.99, 4.07) | 2.20 (1.23, 3.16) | 0.34 (-1.37, 2.04) | 0.7005 |
| *Myalgia - any* | 3.65 (2.11, 5.19) | 3.12 (2.14, 4.10) | 0.52 (-1.21, 2.25) | 0.5527 |
| *Myalgia - moderate or severe* | 3.20 (0.51, 5.90) | 2.17 (0.71, 3.63) | 1.03 (-1.64, 3.70) | 0.4479 |
| *Nausea - any* | 3.31 (1.61, 5.00) | 1.74 (1.17, 2.30) | 1.57 (-0.09, 3.22) | 0.0632 |
| *Chills - any* | 2.10 (1.32, 2.88) | 1.92 (1.07, 2.77) | 0.18 (-0.85, 1.21) | 0.7289 |
| ***L-AEFIs*** |  |  |  |  |
| *Redness - any* | 18.66 (16.76, 20.55) | 5.90 (3.72, 8.09) | 12.76 (9.96, 15.55) | <0.0001 |
| *Redness - moderate or severe* | 7.14 (5.61, 8.66) | 4.24 (2.06, 6.42) | 2.90 (0.41, 5.39) | 0.0226 |
| *Induration - any* | 16.62 (14.51, 18.73) | 5.54 (3.66, 7.41) | 11.09 (8.45, 13.72) | <0.0001 |
| *Induration - moderate or severe* | 5.55 (4.08, 7.02) | 3.23 (2.66, 3.81) | 2.32 (0.75, 3.89) | 0.0037 |
| *Pain - any* | 6.18 (4.66, 7.70) | 4.37 (3.86, 4.88) | 1.81 (0.22, 3.39) | 0.0254 |
| *Pain - moderate or severe* | 4.77 (1.69, 7.85) | 3.13 (2.54, 3.72) | 1.64 (-1.54, 4.82) | 0.3124 |

<sup>&</sup>Average Treatment Effect <sup>\*</sup>weighted for age and HIV status

### Local reactions

LIS-AEFIs were reported by a total of 852 (90.9%) participants, with a higher proportion in those receiving the ID route (94.4% vs 80.0%, p<0.001).

Among LIS-AEFIs, redness and induration at the injection site were the most frequently reported adverse effects (80.9% and 81.0%, respectively), with a significantly higher proportion (about twice as high) in the ID group than in the SC group (p<0.001); a similar difference was also observed considering only moderate or severe grade adverse events.

In contrast, 74.7% of participants in the SC group reported any-grade pain at the injection site vs. 69.2% in the ID group, although there was no evidence for a difference (p=0.14); the same applied when considering moderate or severe grade pain (34.2% vs. 30.1%, p=0.13).

After controlling for HIV and age, we found a higher risk of occurrence for any grade of redness and induration at the local site in the ID than in the SC group (p<0.001). Conversely, a higher degree of pain was reported by participants who received the vaccine through the SC modality (p<0.002). ORs with 95% CI from fitting the multinomial regression models are reported in Table 3. Results were similar after restricting the analysis to the 0-6 days diary data (Supplementary Table 1).

Local redness and induration (regardless of grade) were the most long-lasting symptoms, especially in the ID group (on average, 18.7 and 16.6 days in ID vs. 5.9 and 5.5 in the SC group, respectively). After controlling for HIV status and age, we estimated that the duration of redness (regardless of grade) in participants who received the vaccine in ID modality was 12.8 days (95% CI: 9.96, 15.55; p < 0.0001) longer than that of subjects receiving the SC strategy. Similar results were observed for induration, which lasted on average 11.1 days (95% CI: 8.45, 13.7; p < 0.0001) longer in the ID vs. SC strategy. After restricting the analysis to only moderate or severe grades, we observed a shorter duration of these AEFIs, although still significantly longer in the ID vs. SC group (7.14 vs. 4.24 days for redness, p=0.023 and 5.55 vs. 3.23 days for induration, p=0.004). Overall, local pain (regardless of grade) had a shorter duration, not exceeding 7 days on average, but still with a longer duration in the ID vs. SC group (mean difference 1.8 days (95% CI: 0.2, 3.4; p=0.025) in the unadjusted analysis, which was however largely attenuated after considering only moderate and severe grade and controlling for age and HIV-status (p=0.312, Table 4).

### Immunogenicity

Finally, in a subset of 171 (18%) vaccine recipients for whom samples have been stored and analyzed we compared the average change in immunogenicity one month after the completion of the vaccination cycle according to the route of administration (ID vs SC). Our counterfactual analysis controlling for HIV and age showed some evidence for a larger increase of MPXV-specific IgG (mean diff=0.26 log_2_, p=0.05) and nAb (0.34 log_2_, p=0.08) titers in favour of the ID vs. SC administration. Weaker evidence for a difference by administration route was found in the variation of MVA-BN specific T-cell response measured by Elispot assay (0.41, p=0.18) (Table 5).

**Table 5.** Potential average change one month after the completion of vaccination cycle according to the route of administration of the first dose and ATE^&^ from fitting a linear regression model (log_2_ scale)

|  | **Mean (log<sub>2</sub>)**<br>**in ID**<br>**(95% CI)** | **Mean (log<sub>2</sub>)**<br>**in SC**<br>**(95% CI)** | **ATE\* (95% CI)** | **p-value** |
| --- | --- | --- | --- | --- |
| Elispot | 1.74 (1.31, 2.17) | 1.33 (0.94, 1.73) | 0.41 (-0.19, 1.01) | 0.183 |
| IgG | 1.22 (1.00, 1.43) | 0.96 (0.80, 1.13) | 0.26 (-0.00, 0.51) | 0.053 |
| nAbs | 0.84 (0.53, 1.16) | 0.50 (0.28, 0.72) | 0.34 (-0.04, 0.72) | 0.076 |

<sup>&</sup>Average Treatment Effect; <sup>\*</sup>weighted for age and HIV-status

## Discussion

This work showed that the intradermal route of administration of the MVA-BN vaccine was safe and well tolerated despite higher reactogenicity than the subcutaneous route. We did not observe serious adverse effects or syncope, as recently reported by the manufacturer^11^. However, among the participants receiving the vaccine ID, we observed a slightly higher prevalence of headache and fatigue than in the SC group. This difference, already mild in the unadjusted analysis, was even more mitigated after a multinomial analysis adjusted for age and HIV status. Local redness and induration were confirmed to be more prevalent (94%) and long-lasting (around 18 days) among participants receiving the intradermal route than those receiving the subcutaneous one. On the contrary, pain was more common after subcutaneous administration. These findings are quite expected considering the mode of inoculation and are consistent with other reports. Recently, Frey et al. reported proportions of adverse effects in people receiving the intradermal dose of 97% for the local and 79% for the systemic symptoms, without evidence of severe grade events. Moreover, local symptoms lasted over a month after the intradermal administration^12^.

Although expected and not severe, redness and induration in the forearm were not well accepted by vaccinated people because they were considered a mark of vaccination and, consequently, of sexual behaviour associated with a high risk of contracting mpox.

Stigma could represent a barrier to vaccination, especially in countries where discrimination and racism are deep-rooted, and the LGBT community is criminalized^26^. Anyway, MVA-BN could also be inoculated intradermally into the upper back just below the shoulder blade or into the skin of the shoulder above the deltoid muscle^27^, where the spot is less visible than in the forearm.

Knowledge of these aspects might help clinicians provide more appropriate counselling, make people aware of the course of side effects, and, therefore, make vaccination more acceptable and widespread enough to protect the “core group” that sustains the virus transmission and contributes to its possible spread to the general population^28^.

Regarding immunogenicity, our data collected one month after completing the vaccination cycle found a substantially equivalent immune response between the participants receiving the first dose intradermally and those receiving it subcutaneously. Differently from previously published data, analyzing humoral immune response against vaccinia virus VACV, not MPXV, we measured both titers of specific IgG and nAbs against MPXV, and we found that slightly higher titers of MPXV IgG and nAbs were elicited after the homologous (ID+ID) course of vaccination that after the heterologous (SC+ID) one.

These findings represent the immunological counterpart of a report from May 2022 to May 2024 in the USA, showing a higher incidence of breakthrough infections among people fully vaccinated with the heterologous than the homologous intradermal vaccination cycle. Of note, in our cohort, we did not observe breakthrough infections^29^.

Finally, this is the first analysis providing data regarding the cellular immune response according to the administration route of MVA-BN. There was no evidence for a difference in the cellular immune response between participants receiving the first dose intradermally or subcutaneously.

T-cell response has been known to be crucial for the control and resolution of poxvirus infections^30,31^, and T-cell reactivity to VACV and MPXV was detected decades post-vaccination with first-generation vaccine, suggesting a role of long-lasting cross-reactive T-cell memory responses in vaccine efficacy^32^. A good T-cell response was also demonstrated after third-generation MVA-BN^33^. T-cells elicited from VACV-based vaccines were found to recognize MPXV-derived epitopes, suggesting that it is crucial for the cross-reactivity between different Orthopox strains^34^. Consequently, T-cells could contribute to the recognition of the different MPXV clades and to a broader vaccine efficacy.

Recently, the Afro-Mpox bulletin showed a spreading outbreak involving several African countries, and clade Ib and clade IIb have been sequenced^35^. In the wake of these reactogenicity and immunological data, we support the use of vaccination by MVA-BN administered by an intradermal route in low-income countries, as WHO suggested, even still in the absence of specific data against the clade Ib. Thus, the sparing-dose protocol of ID-based vaccination may provide a cost-effective approach to the current global vaccination campaign.

Some limitations need to be stated. First, the study’s observational design could account for unmeasured bias, although the analysis was controlled for the main confounders. Furthermore, we cannot compare the two homologous cycles (SC+SC vs ID+ID) because the second doses were only administered intradermally due to the regulatory recommendations. For this reason, we have limited the comparison to the first dose. As mentioned, although we extensively assessed the immune response against MPXV clade IIb, we cannot provide information on the humoral and cellular response against Clade I.

In conclusion, our study demonstrated that the intradermal route of administration of MVA-BN elicited slightly higher titers of MPXV-specific IgG and nAbs than the subcutaneous route. At the same time, no differences in cellular response were detected. MVA-BN was globally well tolerated despite higher reactogenicity with the intradermal than the subcutaneous route. Based on these results, we believe that intradermal administration of MVA-BN is feasible, safe, and immunogenic, and we support the use of this dose-sparing strategy to increase the feasibility of a broad vaccination campaign to control the current multiclade mpox outbreak in Africa.

## Data Availability

Data are available upon reasonable request from the corresponding author.

## Acknowledgements

We thank the participants who gave their time to the project, the nurse, the laboratory staff, and the bio-banking personnel. Thanks to all the members of the *Mpox Vaccine Lazio Study Group:* C Aguglia, A Antinori, E Anzalone, A Barca, M Camici, F Cannone, P Caputi, R Casetti, L Caterini, C Cimaglia, E Cimini, F Colavita, L Coppola, R Corso, F Cristofanelli, S Cruciani, N De Marco, G Del Duca, G D’Ettorre, S Di Bari, S Di Giambenedetto, P Faccendini, F Faraglia, D Farinacci, A Faticoni, C Fontana, M Fusto, R Gagliardini, P Gallì, S Gebremeskel, G Giannico, S Gili, E Girardi, G Grassi, MR Iannella, A Junea, D Kontogiannis, A Lamonaca, S Lanini, A Latini, D Lapa, M Lichtner, MG Loira, F Maggi, A Marani, M Marchili, R Marocco, A Masone, C Mastroianni, I Mastrorosa, G Matusali, V Mazzotta, S Meschi, S Minicucci, A Mondi, V Mondillo, A Nappo, G Natalini, E Nicastri, S Notari, A Oliva, A Parisi, J Paulicelli, C Pinnetti, P Piselli, MM Plazzi, A Possi, G Preziosi, R Preziosi, G Prota, M Ridolfi, S Rosati, A Russo, L Sarmati, P Scanzano, L Scorzolini, C Stingone, E Tamburrini, E Tartaglia, V Tomassi, F Vaia, A Vergori, M Vescovo, S Vita, J Volpi, P Zuccalà.

## Authors contribution

VM, AA, and ACL conceived the study; VM, GMa, EC, and FC wrote the protocol. PP and VM wrote the first draft of the manuscript. AA, FM, ACL, EC, GMa, FC, EG, EN, CF and CA revised the manuscript. GMa, FC, SM, LB, SN, RC, and GG performed the laboratory analyses. PP, JP, AC and CC was responsible for data management. PP and ACL performed statistical analysis. VM, RE, AG, GMi, AO, SG, and RG enrolled and followed the patients during the study time points. ML, LS, ET, CM, and AL contributed to the enrollment. PG, AS, FV, and AB contributed to the realization of the study. VM and AA provided the grant for funding the study. All authors gave their final approval of the submitted version.

## Competitive of interests

AA received a grant from Bavarian Nordic for participation in conferences. The other authors declared no conflicts of interest.

## Data sharing statement

Data are available upon reasonable request from the corresponding author.

