## Supplemental Table 1 for "Reactogenicity and immunogenicity against MPXV of the intradermal administration of Modified Vaccinia Ankara compared to the standard subcutaneous route"

**Supplementary Table 1. Prevalence and risk of developing different grades of Systemic and Local Adverse Effects Following Immunisation (S-AEFIs and L-AEFIs) with MVA-BN Vaccine up to six days from fitting a multinomial logistic regression according to administration route - intradermal (ID) vs subcutaneous (SC).**

| | | Prevalence of S-AEFIs and OR (ID vs. SC) up to 6 days from vaccination | | | | | |
|---|---|---|---|---|---|---|---|
| **Systemic symptoms** | **max severity** | **ID n (%)** | **SC n (%)** | **Unadjusted OR (95% CI) ID vs. SC** | **p-value** | **Adjusted* OR (95% CI) ID vs. SC** | **p-value** |
| ***Fatigue*** | None | 401 (56.0) | 137 (60.9) | 1 | 0.001 | 1 | 0.856 |
| | Mild | 174 (24.3) | 35 (15.6) | 1.70 (1.13, 2.56) | | 0.85 (0.37, 1.98) | |
| | Moderate | 106 (14.8) | 29 (12.9) | 1.25 (0.79, 1.97) | | 1.41 (0.54, 3.69) | |
| | Severe | 35 (4.9) | 24 (10.7) | 0.50 (0.29, 0.87) | | 1.20 (0.38, 3.81) | |
| ***Headache*** | None | 500 (69.8) | 166 (73.8) | 1 | 0.022 | 1 | 0.065 |
| | Mild | 152 (21.2) | 29 (12.9) | 1.74 (1.13, 2.69) | | 2.66 (1.13, 6.27) | |
| | Moderate | 47 (6.6) | 22 (9.8) | 0.71 (0.42, 1.21) | | 0.95 (0.25, 3.67) | |
| | Severe | 17 (2.4) | 8 (3.6) | 0.71 (0.30, 1.66) | | 6.60 (0.68, 63.98) | |
| ***Myalgia*** | None | 567 (79.2) | 172 (76.4) | 1 | 0.811 | 1 | 0.970 |
| | Mild | 92 (12.8) | 32 (14.2) | 0.87 (0.56, 1.35) | | 1.03 (0.39, 2.72) | |
| | Moderate | 38 (5.3) | 13 (5.8) | 0.89 (0.46, 1.70) | | 1.11 (0.26, 4.73) | |
| | Severe | 19 (2.7) | 8 (3.6) | 0.72 (0.31, 1.67) | | 1.47 (0.31, 6.94) | |
| ***Nausea*** | None | 646 (90.2) | 199 (88.4) | 1 | 0.176 | 1 | 0.895 |
| | Mild | 49 (6.8) | 15 (6.7) | 1.01 (0.55, 1.83) | | 1.20 (0.31, 4.71) | |
| | Moderate | 11 (1.5) | 9 (4.0) | 0.38 (0.15, 0.92) | | 0.53 (0.09, 3.02) | |
| | Severe | 10 (1.4) | 2 (0.9) | Nd | | Nd | |
| ***Chills*** | None | 656 (91.6) | 202 (89.8) | 1 | 0.260 | 1 | 0.803 |
| | Mild | 35 (4.9) | 15 (6.7) | 0.72 (0.38, 1.34) | | 0.89 (0.25, 3.09) | |
| | Moderate | 13 (1.8) | 7 (3.1) | 0.57 (0.23, 1.45) | | 0.31 (0.03, 3.17) | |
| | Severe | 12 (1.7) | 1 (0.4) | Nd | | Nd | |
| ***Vomit*** | None | 710 (99.2) | 219 (97.3) | 1 | 0.559 | 1 | 0.993 |
| | Mild | 3 (0.4) | 3 (1.3) | 0.31 (0.06, 1.54) | | Nd | |
| | Moderate | 0 (0.0) | 3 (1.3) | Nd | | Nd | |
| | Severe | 3 (0.4) | 0 (0.0) | Nd | | Nd | |

| | | Prevalence of L-AEFIs and OR (ID vs. SC) | | | | | |
|---|---|---|---|---|---|---|---|
| **Local symptoms** | **max severity** | **ID n (%)** | **SC n (%)** | **Unadjusted OR ID vs. SC** | **p-value** | **Adjusted* OR ID vs. SC** | **p-value** |
| ***Redness*** | None | 46 (6.5) | 137 (60.9) | 1 | <.001 | 1 | <0.001 |
| | Mild | 168 (23.6) | 43 (19.1) | 11.63 (7.25, 18.67) | | 20.84 (6.05, 71.73) | |
| | Moderate | 325 (45.6) | 25 (11.1) | 38.72 (22.87, 65.54) | | 47.01 (13.15, 168.0) | |
| | Severe | 173 (24.3) | 20 (8.9) | 25.76 (14.56, 45.59) | | 36.37 (9.90, 133.7) | |
| ***Induration*** | None | 67 (9.4) | 121 (53.8) | 1 | <.001 | 1 | <0.001 |
| | Mild | 264 (37.1) | 51 (22.7) | 9.35 (6.12, 14.27) | | 10.84 (4.11, 28.57) | |
| | Moderate | 281 (39.5) | 36 (16.0) | 14.10 (8.92, 22.28) | | 12.95 (4.55, 36.83) | |
| | Severe | 100 (14.0) | 17 (7.6) | 10.62 (5.86, 19.25) | | 14.20 (4.28, 47.14) | |
| ***Pain*** | None | 257 (36.1) | 60 (26.7) | 1 | 0.057 | 1 | <0.001 |
| | Mild | 266 (37.4) | 90 (40.0) | 0.69 (0.48, 1.00) | | 0.21 (0.10, 0.48) | |
| | Moderate | 147 (20.6) | 58 (25.8) | 0.59 (0.39, 0.90) | | 0.09 (0.03, 0.26) | |
| | Severe | 42 (5.9) | 17 (7.6) | 0.58 (0.31, 1.08) | | 0.37 (0.09, 1.45) | |

* Adjusted for age and HIV status; 95 CI: 95% Confidence Interval; Nd: not determined
