## Supplemental Figure S1 for "Reactogenicity and immunogenicity against MPXV of the intradermal administration of Modified Vaccinia Ankara compared to the standard subcutaneous route"

**Supplementary Figure S1. Heatmap summarizing grade and duration of Systemic (Panel a) and Local Injection Site (Panel b) Adverse Effect Following Immunisation (S-AEFI and LIS-AEFI) with MVA-BN Vaccine within 28-days from vaccination according to route of vaccination [Sub-cutaneous (SC): N=225; Intra-dermal (ID): N=718]. Every row represents results for a participant between Days 1-28 which are presented as columns. Cells are color-coded by severity.**

**Panel a**

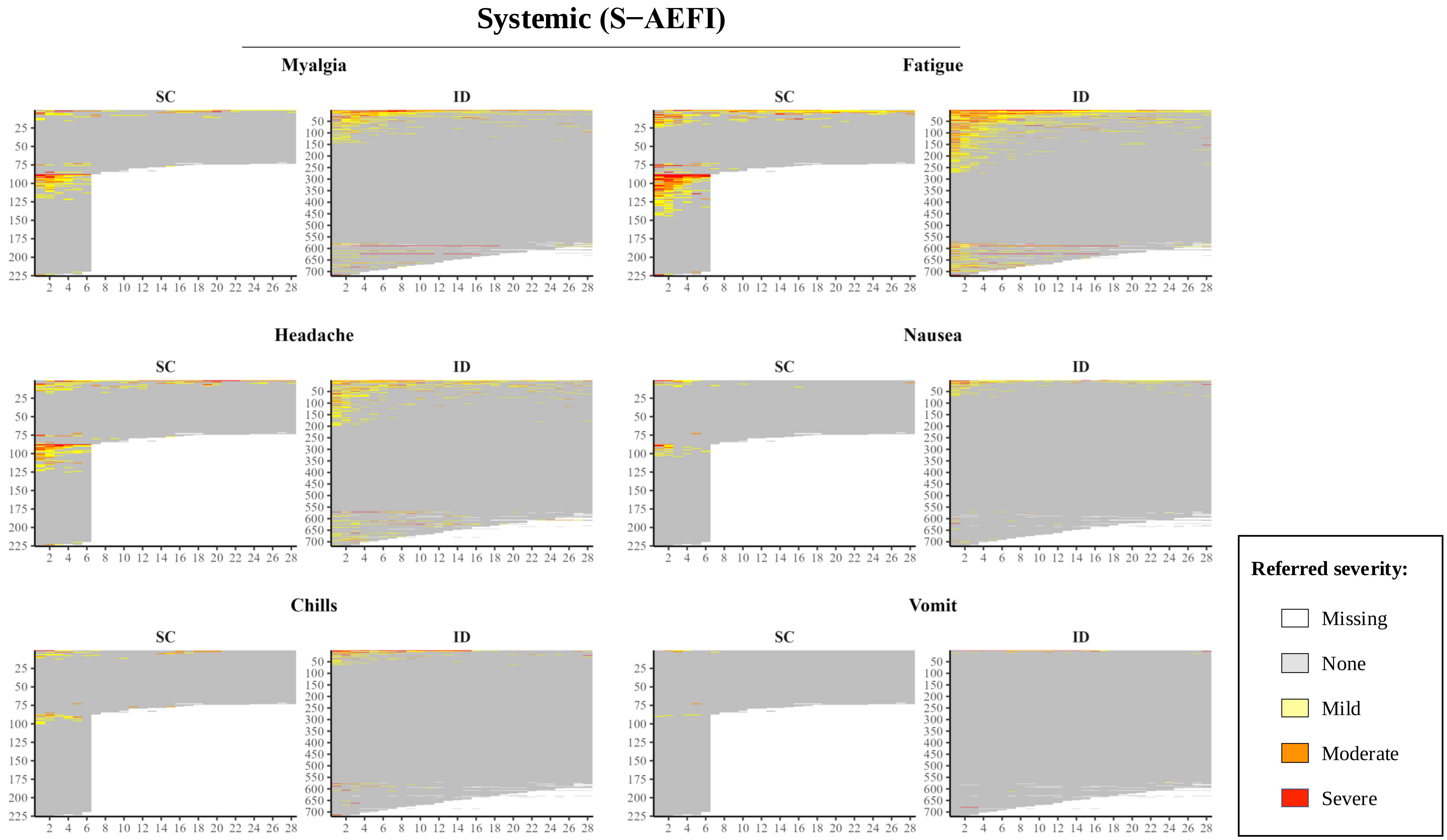

Systemic (S−AEFI)
Myalgia
Fatigue
Headache
Nausea
Chills
Vomit
SC
ID
Referred severity:
Missing
None
Mild
Moderate
Severe

**Panel b**

### Local Injection Site (LIS−AEFI)

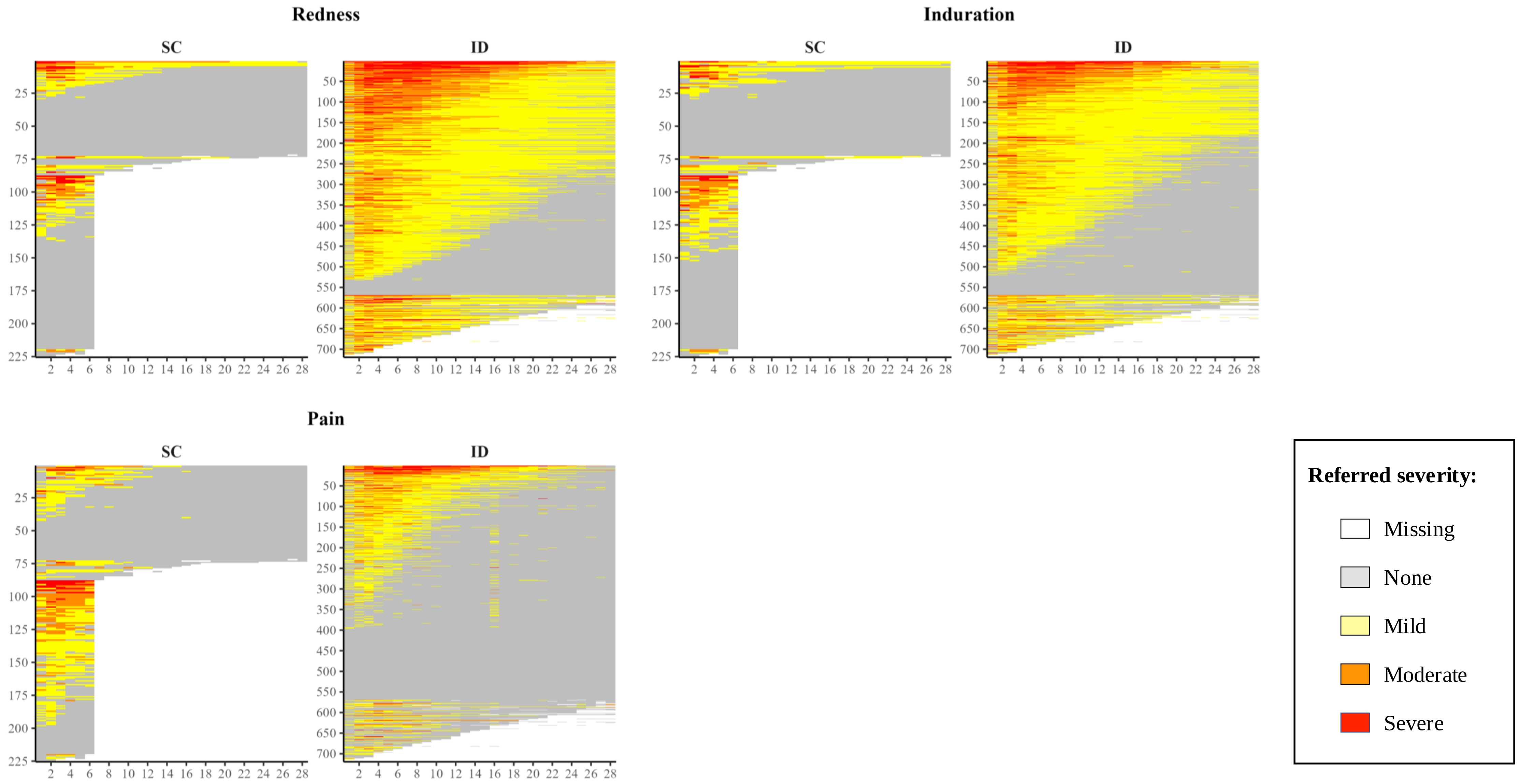
